# Temporal changes in the risk of superspreading events of coronavirus disease 2019

**DOI:** 10.1101/2021.05.10.21256927

**Authors:** Jun-Sik Lim, Eunbi Noh, Eunha Shim, Sukhyun Ryu

## Abstract

In order to identify the temporal change in the possible risk of superspreading events (SSE), we estimated the overdispersion parameter in two different periods of COVID-19 pandemic. We identified the possible risk of SSE was reduced 34% during the second epidemic period in South Korea.

## Introduction

Individual-based interventions, including active contact tracing and isolation, interrupt the transmission chain and reduce the infectiousness of severe acute respiratory syndrome coronavirus-2 (SARS-CoV-2) [1, 2]. To identify changes in the transmissibility of SARS-CoV-2, the daily effective reproductive number has been estimated. However, this estimate has limited interpretability, as it does not reflect the individual variation of infectiousness [3], which is known as overdispersion in the distribution of the number of secondary transmissions and is the deterministic factor of superspreading events (SSE) [4].

The *k* is assumed to be a fixed characteristic of infection, and there is lack of study demonstrating its temporal changes on different epidemic periods. Here, we examined contact tracing data in South Korea to assess temporal changes in the individual variation of the risk of SSE on SARS-CoV-2 transmission

## Methods

We retrieved publicly available data on coronavirus disease 2019 (COVID-19) cases from local public health authorities in South Korea. The authorities provide daily updates of all COVID-19 cases confirmed by consistent real-time reverse transcriptase-polymerase chain reaction (RT-PCR). The data comprised the date of illness onset, date of laboratory diagnosis, and infection source. We excluded the Daegu-Gyeongsangbuk region, where the data were not publicly reported.

Based on the epidemic curve of two epidemic waves in South Korea [5], we divided the study period into two, based on the date of symptom onset of the infector in pairs: Period-1, January 19–April 19, 2020; and Period-2, April 20–October 16, 2020. Furthermore, based on the infection source, we built infectee-infector pairs to estimate the offspring distribution.

Offspring distribution was estimated from the infectee-infector pairs to demonstrate the overdispersion of individual heterogeneity in transmission for each epidemic period. Using the observed distribution of the secondary case, we fitted the negative binomial offspring distribution (Supplementary appendix). This distribution was parameterized with the basic reproductive number (*R*_0_) and dispersion parameter (*k*), which indicate the mean number of secondary cases generated by an index case and the individual level of heterogeneity in transmission, respectively [6, 7]. A smaller k indicates a longer tail of the negative binomial distribution, which means the individual level of variation in secondary cases is higher, and hence the epidemic includes a higher likelihood of SSE [3]. This distribution has been widely used to identify the individual level of variation in infectiousness [6-8]. Furthermore, we estimated the expected proportion of cases responsible for 80% of the total secondary transmission, and the probability of SSE using the estimated *R*_*i*_ and *k*_*i*_, which was taken from previous studies [4, 8] and the probabilities that an index case of SARS-CoV-2 infection results in a cluster of 10 cases or more were estimated (Supplementary appendix). We used the *runjags* package for Markov chain Monte Carlo simulation. All statistical analyses were performed in R version 3.6.1 (R Foundation for Statistical Computing)

## Results

In this study, we identified 675 transmission pairs (104 for Period-1 and 571 for Period-2) with 1070 sporadic cases not linked with other cases (240 for Period-1 and 830 for Period-2). Our estimates suggested that the *k* in Period-1 (0.23, 95% credible interval (CrI) 0.15–0.28) was lower than Period-2 (0.68, 95% CrI: 0.62–0.74) (Figure 1). Furthermore, the expected proportion of cases responsible for 80% of secondary transmission was 14.70% (95% CrI, 12.21–17.36%) in Period-1 and 25.72% (95% CrI, 24.02–27.41%) in Period-2. The probability of SSE was 2.43% (95% CrI: 1.39–4.04%) in Period-1 and 0.24% (95% CrI: 0.14–0.40%). The probability that an index case of SARS-CoV-2 infection results in a cluster of 10 cases or more, was 7.46% (95% CrI, 4.47–11.65%) in Period-1 and 5.74% (95% CrI, 4.24–7.56%) in Period-2 (Figure 2).

**Figure 1.**
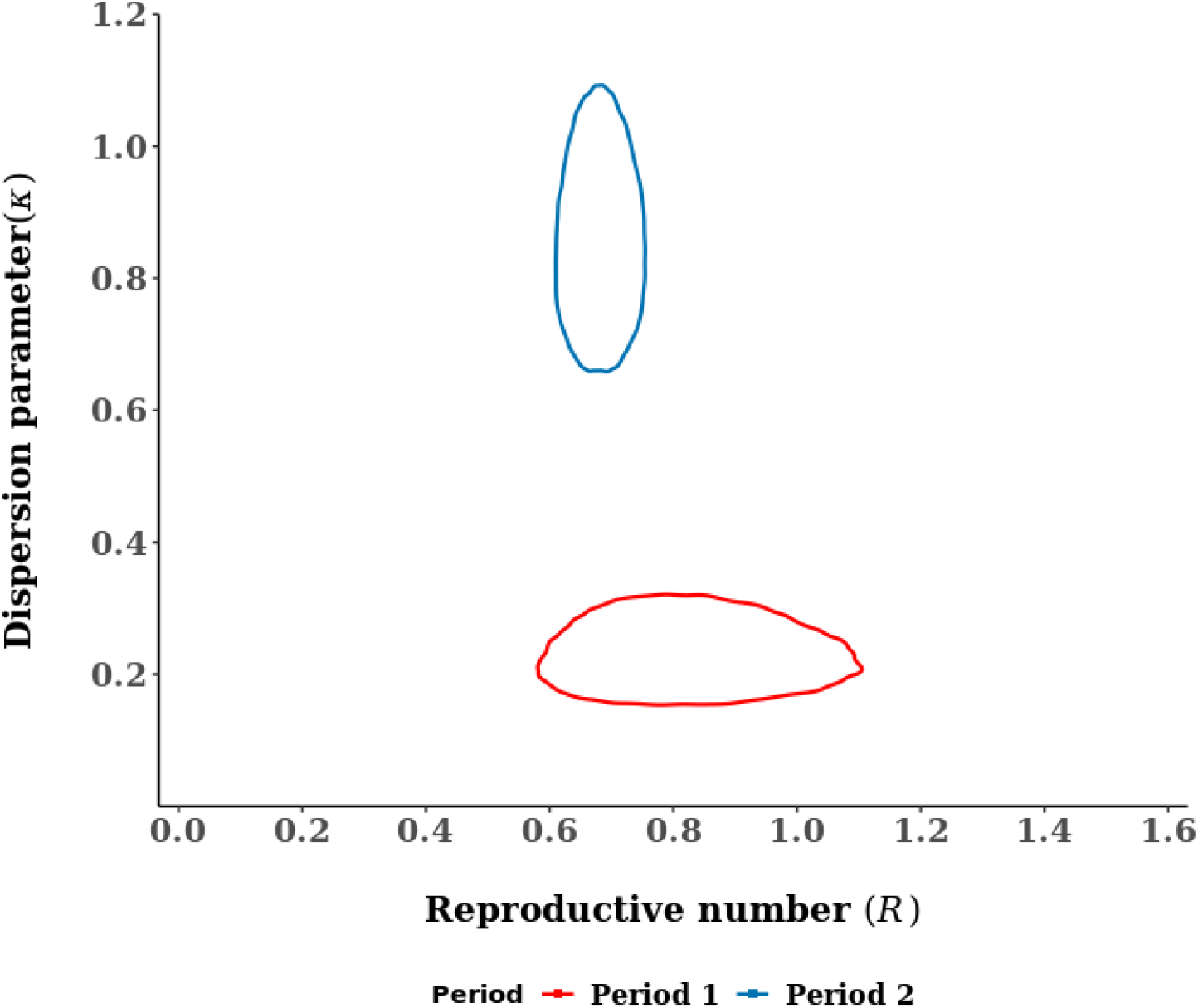
Joint estimates of overdispersion parameter and reproduction number of coronavirus disease 2019. The red line indicates the estimated overdispersion parameter (*k*) in the first period (19 January–19 April 2020). The blue line indicates the estimated *k* in the second period (20 April–16 October 2020). The lines show 95% credible intervals.

**Figure 2.**
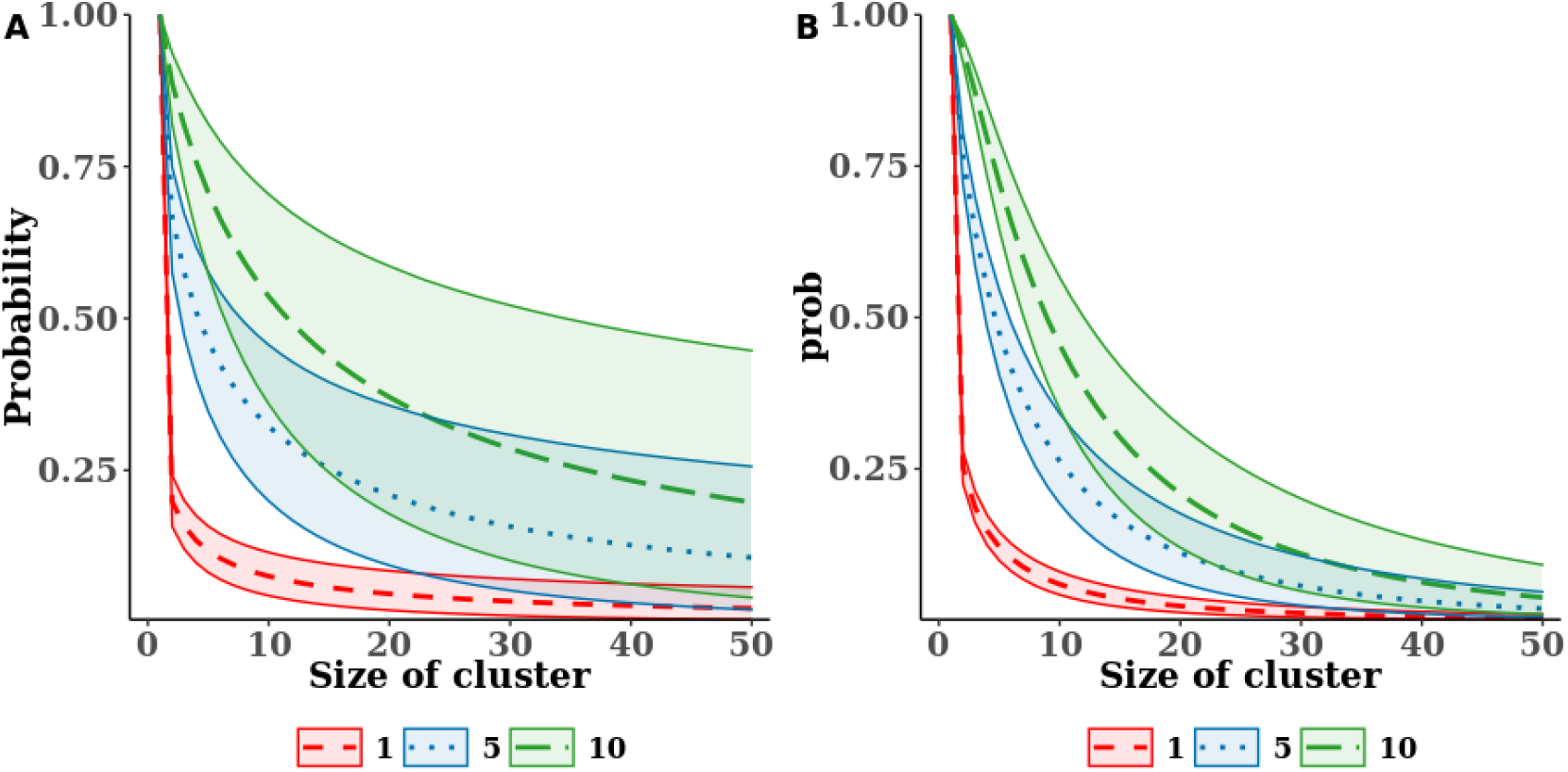
The expected probabilities that the different number of index case generates clusters of given size. Dashed, dotted, and long-dashed line indicate the median estimates of the expected probabilities of one, five, and ten infectors generate clusters of given size, respectively. The green, red, and blue areas covered by solid lines indicate 95% credible intervals.

## Discussion

Our findings suggest that the individual variation of SARS-CoV-2 infectiousness is highly over-dispersed, which is corroborated by previous reports that *k* was estimated to be 0.43 in Hong Kong [8]. Furthermore, our results showed that *k* was larger in the latter epidemic period, which is consistent with a previous study that the serial interval was reduced over time as enhanced nonpharmaceutical intervention (NPI) was implemented in the community [2]. This reduction suggested that the risk of SSE was lower in the latter period of the epidemic. This may affected by improved contact tracing with the digital QR code entry logs at public facilities began June 10, 2020 to quickly identify the possible COVID-19 case [9] and enhanced social distancing in publicly used facilities (a ban on religious gatherings began August 19, 2020) during Period-2 [10]. Other factors including public’s behavior changes derived by improved risk-awareness of COVID-19 and climatological factor could also influence the transmission dynamics of SARS-CoV-2 and the changes of the risk of SSE.

The South Korean public health authority has modified the level of nationwide nonpharmaceutical intervention, primarily using data on the mobility of people, the number of COVID-19 cases, the number of clusters, and the effective reproductive number [11]. Because few cases could account for many COVID-19 cases, monitoring the overdispersion parameter would help to provide relevant information on transmission dynamics of SARS-CoV-2 to the public health authority.

There are several limitations in our study. First, we used a case report from public health authorities that is not free from under-reporting of cases and could bias our results. Therefore, our result of the possible risk of SSE is likely to be a lower bound estimate. Second, we have not included cases from the Daegu-Gyeongsangbuk region, which originated primarily from the Shincheonji religious group. A previous report demonstrated that there was a delay of several days in detecting cases that could affect the size of the cluster. Therefore, this group may not reflect the typical characteristics of the transmission. Third, we have not included the latter period of COVID-19 pandemic which the number of COVID-19 case was larger than previous two period in the present study. However, this outbreak was mainly originated anti-governmental rallies related with religious groups which avoiding contact tracing were reported [12]; hence, did not reflect characteristics of typical community transmission in South Korea. Fourth, we had not included the genetic sequencing data of the cases, as genetic sequencing was not conducted in most cases. Therefore, we could not rule out a pseudo-outbreak of COVID-19 in which the PCR results were false-positive [13] or infection was acquired from outside the cluster.

In conclusion, our study suggests that there were temporal changes in the possible risk of SSE in South Korea. Ongoing monitoring of the risk of SSE would help to assess the impact of public health measures implemented in the community and to provide data for policy decision-making.

## Data Availability

The data can be retrieved from local public health authorities in South Korea.

## Ethical approval

Not applicable. The use of the publicly available dataset did not need ethical approval.

