## Supplementary material for "Temporal changes in the risk of superspreading events of coronavirus disease 2019"

### Technical Appendix: Temporal changes of the risk of superspreading events of coronavirus diseases 2019

**Affiliations:**
1. Graduate School of Public Health, Seoul National University, Seoul, 08826, Republic of Korea

2. College of Veterinary Medicine and Institute of Veterinary Science, Kangwon National University, Chuncheon, Korea

3. Department of Preventive Medicine, Konyang University College of Medicine, Daejeon, 35365, Republic of Korea

4. Department of Mathematics, Soongsil University, Seoul, 06978, Republic of Korea

### Author for correspondence: Prof. Sukhyun Ryu

Department of Preventive Medicine, Konyang University College of Medicine, R707, Myungok-Euihak Gwan, 158, Gwanjeodong-ro, Seogu, Daejeon, South Korea.

**Word count (abstract):** 42/50; **Word count (main text):** 1054/1500

Running title: Change in the risk of superspreading of COVID-19

### Method

Based on the regression model, the probability that an index case ($i)$ for period ($j$) generate the number of secondary case ($y_{i,j}$) is given by

$$Pr\left( Y = y_{i,j} \right) = \frac{\Gamma\left( k_{j} +y_{i,j} \right)}{y_{i,j}!\Gamma\left( k_{j} \right)}\left( \frac{k_{j}}{k_{j} +R_{j}} \right)^{k_{j}}\left( \frac{R_{j}}{k_{j}+R_{j}} \right)^{y_{i,j}}$$

where $k_{j}$ and $R_{j}$ is the estimated dispersion parameter and reproduction number for period $j (j=1 or 2)$.

This model incorporates the covariates into the $k_{j}$ and $R_{j}$ as log-linear model like follows:

$$\log\left( R_{j} \right)=\alpha_{R}+\sum\beta_{R}X_{R}$$

$$\log\left( k_{j} \right)=\alpha_{k}+\sum\beta_{k}X_{k}$$

where $\alpha_{R_{0}}$ and $\alpha_{k}$ are intercepts, $\beta_{R}$ and $\beta_{k}$ are vectors of coefficients of period variable, $X_{R}$ and $X_{k}$ are vectors of period variable for $k_{j}$ and $R_{j}$, respectively.

To understand the transmission dynamics of COVID-19 on different two epidemic period, two analyses were conducted: (1) analysis for only period variable, (2) analysis for period variable adjusted with the sex and age of infector. In analysis for only period variable, only period variable was incorporated in both $k_{j}$ and $R_{j}$ parts of negative binomial regression; In analysis for period variable adjusted with the sex and age of infector, sex and age of infector as well as period variable was incorporated in both $k_{j}$ and $R_{j}$ parts of negative binomial regression.

Given the estimated reproduction number and dispersion parameter for each period, the proportion of infected person responsible for 80% of secondary cases $P_{80\%}$ was calculated using equations from the previous studies [1-3]. The proportion $P_{80\%}$ is given by

$$1-P_{80\%}=\int_{0}^{X} NB\left( \left\lfloor x \right\rfloor;k_{j}, \frac{k_{j}}{R_{j}+ k_{j}} \right)dx$$

where $X$ satisfies

$$1-0.20=\frac{1}{R_{j}}\int_{0}^{X} \left\lfloor x \right\rfloor NB\left( \left\lfloor x \right\rfloor;k_{j}, \frac{k_{j}}{R_{j}+ k_{j}} \right)dx$$

Moreover, the risk of being infector $Prob\left( infector | infection \right)$and non-infector

Furthermore, with the threshold for SSE as 6 secondary cases defined in the previous study [1], the proportion of SSE was estimated for each period with the equations like below :

$$\int_{6}^{\infty} NB\left( \left\lfloor x \right\rfloor;k_{j}, \frac{k_{j}}{R_{j}+ k_{j}} \right)dx=1- \int_{0}^{6} NB\left( \left\lfloor x \right\rfloor;k_{j}, \frac{k_{j}}{R_{j}+ k_{j}} \right)dx$$

Finally, using the branching process, the expected probability that one index case of SARS-CoV-2 infection results in a cluster of size $s$ were estimated with the equations from previous studies [2, 3] given by

$$r_{s}= \frac{\Gamma\left( k_{j}s +s-1 \right)}{\Gamma\left( k_{j}s \right)\Gamma\left( s+1 \right)}\left( \frac{R_{j}}{k_{j}} \right)^{s-1}\left( \frac{k_{j}}{k_{j}+R_{j}} \right)^{k_{j}s +s-1}$$

where the probability of cluster of size $s$ or greater could be estimated like below

$$p_{s}=1-\sum_{l=1}^{s-1} r_{l}$$

Following the condition that$N$ seed cases were introduced into the totally susceptible populations, the estimated probability that at least one cluster of size $s$ or greater occurs is

$$P_{N,s}= 1- {(1-p_{s})}^{N}$$

Note that $P_{N,s}$ is the same with $p_{s}$ with the condition that one index case was introduced ($N=1$).

We used a Bayesian Markov Chain Monte Carlo (MCMC) sampling. Four chains of 50,000 iterations were obtained with 20,000 burn-in. Prior distribution for the coefficients in each regression were set as normal distribution with mean of 0 and variance of 100,000. Convergence was checked by the trace plot visually and Gelman-Rubin-Brooks diagnostic [4]. The posterior distribution of the estimates is demonstrated with the median and the 95% credible intervals (95% CrI).
